# EVALUATION OF A RAPID ANTIGEN TEST FOR SARS-COV-2 IN SYMPTOMATIC PATIENTS AND THEIR CONTACTS: A MULTICENTER STUDY

**DOI:** 10.1101/2021.05.24.21257020

**Authors:** Ireri Thirion-Romero, Selene Guerrero-Zúñiga, Alexandra Arias-Mendoza, Dora Patricia Cornejo-TJuárez, Patricia Meza-Meneses, Darwin Stalin Torres-Erazo, Thierry Hernández, Arturo Galindo-Fraga, Isabel Villegas-Mota, Jesús Sepúlveda-Delgado, Santiago Ávila Ríos, Eduardo Becerril-Vargas, Rosario Fernández-Plata, TIT Midori Pérez-Kawabe, Joel Armando Vázquez Pérez, Simón Kawa Karasik, Gustavo Reyes Terán, José Rogelio Pérez-Padilla, Rapid COVID-19 Antigen Test Group

## Abstract

**BACKGROUND:** Point-of-care rapid tests to identify SARS-CoV-2 can be of great help because, in principle, they allow decisions to be made at the site of care for treatment, or for the separation of cohorts avoiding cross-infection, especially in emergency situations.

**METHODS:** A cross sectional study in adults requesting care in Emergency Rooms (ER), or the outpatient clinics of referral hospitals for COVID-19, to define the diagnostic characteristics of a rapid antigen test for SARS-CoV-2 (the Abbott Panbio™) having as a gold standard the RT-PCR for SARS-CoV-2. Health personnel in a routine situation within an active pandemic in several cities of Mexico performed the tests.

**RESULTS:** A total of 1,069 participants with a mean age of 47 years (SD 16 years), 47% with a self-reported comorbidity, and an overall prevalence of a positive RT-PCR test of 45%, were recruited from eight hospitals in Mexico. Overall sensitivity of the Panbio test was 54.4% (95%CI 51-57) with a positive likelihood ratio of 35.7, a negative likelihood ratio of 0.46 and a Receiver-Operating Characteristics curve area of 0.77.

Positivity for the rapid test depended strongly on an estimate of the viral load (Cycle threshold of RT-PCR, Ct), and the days of symptoms. With a Ct≤25, sensitivity of the rapid test was 0.82 (95%CI, 0.76-0.87). For patients during the first week of symptoms sensitivity was 69.6% (95%CI 66-73). On the other hand, specificity of the rapid test was above 97.8% in all groups.

**CONCLUSIONS:** The Panbio™ rapid antigen test for SARS-CoV-2 has a good specificity, but due to low and heterogeneous sensitivity in real life, a negative test in a person with suggestive symptoms at a time of community transmission of SARS-CoV-2 requires confirmation with RT-PCR, and after the first week of symptoms, sensitivity decreases considerably.

## INTRODUCTION

Rapid tests to identify infectious agents can be highly useful since, in principle, they allow decisions to be made at the site of care (Point-of-Care, POC) for treatment selection, or for the separation of cohorts to avoid cross-infection. Rapid tests are especially useful in emergency situations, such as those currently being experienced in the context of the COronaVIrus Disease 19 (COVID-19) pandemic. For influenza and other respiratory viruses, rapid tests are readily available and have shown clinical benefits (1-4), although not in all evaluations (5). Rapid tests, at POC can be employed for screening purposes in asymptomatic people, for diagnostic purposes in persons with symptoms suggestive of the disease, or for contact tracing and epidemiological purposes in persons who had contact with suspect or confirmed cases. These situations in which the pre-test probabilities of SARS-CoV-2 infection are very different, lead to different demands (6). A recent Cochrane review showed the urgent need for prospective and comparative evaluation of rapid tests in the context of COVID-19 (7). Having a reliable rapid test would be highly desirable, especially in places with poor infrastructure or without easy access to standard laboratory tests, but also at reference sites, especially when faced with the possible arrival of patients with similar clinical manifestations but with infections with different virus (8). Rapid tests could also be performed on the same subject on several occasions at low cost, for the purposes of detection and isolation of positive cases and for epidemiological surveillance, even when these usually have lower sensitivity than Reverse Transcription Polymerase Chain Reaction (RT-PCR)-based tests (9).

In the context of the COVID-19 pandemic, several international regulatory bodies have granted authorization for the emergency use of rapid tests for the presumptive diagnosis of Severe Acute Respiratory Syndrome CoronaVirus 2 (SARS-CoV-2) infection based on the identification of antigens. While the overall recommendation is to confirm results with tests currently considered as gold standard, such as PCR-based tests, a readily available result obtained with a rapid antigen test can aid in making several important decisions in the clinical-care workflow.

Here, we assessed the performance of a rapid antigen test for SARS-CoV-2 as a diagnostic tool in symptomatic patients who arrived at the Emergency Rooms (ER) and outpatient clinics of referral hospitals for COVID-19, under real working conditions, by health personnel, in the midst of the pandemic and also in symptomatic or asymptomatic contacts of patients diagnosed with SARS-CoV-2.

## METHODS

This is an observational, cross-sectional study performed in eight tertiary-care referral hospitals for COVID-19 in Mexico, comparing the performance of a rapid antigen test, against the gold standard, the RT-PCR test. The participating institutions were part of the Mexican National Institutes of Health (NIH) network, including the National Institute of Respiratory Diseases (INER), the National Institute of Cancer (INCAN), the National Institute of Cardiology (INCIC), the High Specialty Regional Hospital Ciudad Salud in Tapachula Chiapas, the High Specialty Regional Hospital of Mérida Yucatán (HRAE Mérida), the High Specialty Regional Hospital in Ixtapaluca State of Mexico (HRAE Ixtapaluca), the National Institute of Medical Sciences and Nutrition (INCMNSZ), and the National Institute of Perinatology (INPER). The protocol was revised and approved by a single Institutional Review Board designated by the NIH authority for this study.

### Inclusion criteria

Patients ≥18 years of age, who presented at the emergency services or screening sites of the participating hospitals, with respiratory symptoms consistent with COVID-19 or influenza syndrome, and who provided written informed consent to participate in the study, were included, regardless of hospitalization status. Contacts of confirmed COVID-19 cases, who presented to the same sites for evaluation (mostly with respiratory symptoms but some asymptomatic), were also enrolled. Participants were recruited on weekdays, during the morning-afternoon shift, from the eight participating institutions.

### Gold standard

For the purpose of this work, the “Berlin” SARS-CoV-2 RT-PCR methodology recommended by the World Health Organization (WHO) (10), was considered the gold standard. All participating institutions implemented this test *in situ* and were accredited by the corresponding national authority, the National Epidemiological Reference Institute (InDRE) based on the detection of 4 SARS-CoV-2 markers: the N, E, ORF and RdRp genes. The RT-PCR test for SARS-CoV-2 was performed according to the Berlin protocol, on nasopharyngeal swab samples. In all cases Cycle treshold (Ct) for the different gene targets were requested. Laboratory personnel were blinded to results for the rapid tests.

#### Procedures in the Emergency Rooms, in outpatient clinics and in contacts

After explaining what the test consists of and the potential participant providing their written informed consent, the rapid test and RT-PCR were performed in the Emergency Room (ER), in triage, or at the usual reception site for probable patients with COVID-19.

### Rapid SARS-CoV-2 Antigen Test

The Panbio™ COVID-19 Ag Rapid Test Device (nasopharyngeal) (Abbott Diagnostics Korea, Inc. Ref. 41FK10) was evaluated. This test does not require additional equipment and is approved by the corresponding regulatory agencies in the United States, Europe, and Mexico. The manufacturer reports a sensitivity of 93.3% (95% CI of 83-98%), and a specificity of 99.4% (95% CI, 95-99.3%) with a detection limit of 1.5×10^1.8^ median Tissue Culture Infectious Dose (TCID_50_)/mL (11).

The tests were carried out by healthcare personnel at the ER or the triage area, following the manufacturer’s instructions. All participating healthcare personnel was specifically trained in obtaining nasopharyngeal swabs and in the use of the rapid test. In all cases, a stopwatch was utilized to record the exact time of the reading. The result and a photograph of the cassette were incorporated into the database for control and later verification if necessary. Clinical information was retrieved using a REDCap database (REDCap 10.31-2021), including the WHO COVID-19 severity classification (12), the use of respiratory support (oxygen by nasal prongs or high flow), mechanical ventilation, and the presence and duration of respiratory symptoms. For hospitalized patients, the results of routine laboratory tests and clinical data were subsequently collected, including a complete blood count, serum electrolytes, glucose, urea, creatinine, ElectroCardioGram (ECG), liver function tests, urinalysis, and Chest Computed Tomography.

A sample number of 600 subjects, with at least 300 symptomatic patients, and 300 contacts was considered in the protocol (13, 14), allowing for a study power of 0.8 even with a prevalence of positivity for the test of 0.2, and a sensitivity and specificity of 0.8.

#### Blinding

The laboratory that processes the RT-PCRs and the users of the rapid tests did not know the results of the other tests.

## STATISTICAL ANALYSIS

The basic comparison was performed in a 2×2 table between the positive and negative tests by the gold standard and between the positive and negative tests by the rapid test, with which sensitivity, specificity, and positive and negative predictive value, as in a diagnostic test with an existing gold standard. We also evaluated concordance (kappa statistic) between gold standard and the Panbio™ tests.

Patients and contacts and participating hospitals were considered as subgroups. The impact of the duration of symptoms in terms of the positivity of the rapid test was assessed with the positivity to the test as a dependent variable as a function of the duration of symptoms, taking into account the Cycles for the positivity of the test (Ct) and not only a dicothomic positivity or negativity, whose criteria can vary. Age, sex, days from the onset of symptoms (if they occurred), the symptoms that occurred, disease severity based primarily on the type of support needed, time of arrival at the ER, time of the test, time of the result, day and the time of obtaining the PCR result were taken as covariates on order to observe modifying effects on sensitivity and specificity.

Analysis was performed with STATA v13.0 statistical software, with summary statistics for diagnostic tests performed by DISGT and DIAGTEST procedures. The MIDAS procedure (Meta-analytical Integration of Diagnostic Accuracy Studies) was utilized to analyze diagnostic performance across recruiting sites.

## RESULTS

A total of 1,073 participants had a rapid test result, and 1,070 a RT-PCR test result. A total of 1,069 participants (605 women and 464 men; mean age 47**±**16 years) simultaneously had the rapid test and the RT-PCR test result and were further analyzed as follows: 379 were recruited at INER; 250 at INCAN; 32 at INCIC; 41 at Ciudad Salud Tapachula Chiapas; 44 at HRAE Mérida; 80 at HRAE Ixtapaluca; 150 at INCMNSZ; and 93 at INPER. Ct values were reported for 389 out of 477 positive RT-PCR tests (81.5%), given that some of the participating hospitals, mainly outside of Mexico City, required sending the samples for processing in state laboratories, and for analysis we utilized the lowest Ct of those reported for different genes.

**Table 1** describes the clinical characteristics of the study participants. From a total of 1,069; 939 participants had any respiratory symptom (87.8%), and 130 (12.2%) were asymptomatic (usually contacts of a positive relative or coworker). Among participants that reported any symptom in 78% these had lasted for less than 1 week.

**TABLE 1:** Principal characteristics of participants (Means and SD or percentage and N)

|  | Total | Female | Male |
| --- | --- | --- | --- |
|  | (1069) | (605) | (464) |
| Age | 47 (16) | 46 (16) | 46 (17) |
| Days of symptoms | 6 (8) | 6 (10) | 6 (5) |
| Asymptomatic contacts | 12% (130) | 13% (77) | 12% (53) |
| Fever | 33% (356) | 29% (174) | 39% (182) |
| Dry Cough | 37% (397) | 38% (231) | 36% (166) |
| Cough and phlegm | 11% (114) | 10% (58) | 12% (56) |
| Phlegm | 9% (98) | 10% (58) | 9% (40) |
| Dyspnea | 24% (261) | 22% (132) | 28% (129) |
| Any comorbidity | 47% (493) | 48% (279) | 47% (214) |
| Diabetes | 14% (150) | 14% (83) | 14% (67) |
| Hypertension | 19% (198) | 20% (118) | 17% (80) |
| Obesity | 10% (107) | 10% (62) | 10% (45) |
| Any cancer | 10% (105) | 11% (66) | 8% (39) |
| Smoking history | 27% (286) | 20% (123) | 35% (163) |
| Current Tobacco smoker | 10% (108) | 7% (44) | 14% (64) |
| Former smoker | 18% (178) | 14% (79) | 25% (99) |
| Cigarettes per day in smokers | 4(7) | 3(4) | 5(8) |
| Influenza vaccine 2020-2021 | 49% (517) | 51% (306) | 46% (211) |
| COVID vaccine | 5% (49) | 5% (28) | 5% (21) |
| SpO2% | 90(10) | 91(10) | 89(10) |
| Breathing frequency | 21(6) | 21(5) | 22 (7) |
| Oxygen use in the moment of the test | 18% (194) | 14% (82) | 24% (112) |
| ER visit | 57% (606) | 59% (354) | 54% (252) |
| Outpatient clinic | 34% (367) | 34% (203) | 35% (164) |
| Hospitalized | 9% (96) | 8% (48) | 10% (48) |
| Ambulatory | 73% (775) | 74% (449) | 70% (326) |
| Re-admission | 4% (17) | 4% (11) | 3% (6) |
| Positive RT-PCR test | 45%(477) | 41% (251) | 49% (226) |

Percentage (%) or mean (standard deviation) are shown. ER= emergency room,

About half (57%) of the participants requested attention in the ER, and 34% at an outpatient service. After initial screening 73% of all the participants were managed as outpatients. At the time of recruitment only 49 participants (5%) had received one dose of the SARS-CoV-2 vaccine (none had received a full vaccination scheme) and 47% of participants presented at least one comorbidity as follows: 19% had hypertension;14% had diabetes; 10% were obese, and 10% were current smokers.

Considering all of the participants, 45% were positive in the RT-PCR test, and 26% were positive in the rapid test. Positivity of the rapid test was strongly associated with Ct (a surrogate for viral load) (**Figure 1)**, (Table S1) with a sensitivity of 0.82 (95%CI, 0.76-0.87) with a Ct≤25 (**Figure 2)**. Positivity also depended on the days since the onset of symptoms (**Figure 3**) (Table S2), with an initial sensitivity of the rapid test of 0.70 (95%CI 0.63-0.74) during the first week of symptoms, exhibiting a progressive decline.

**FIGURE 1:**
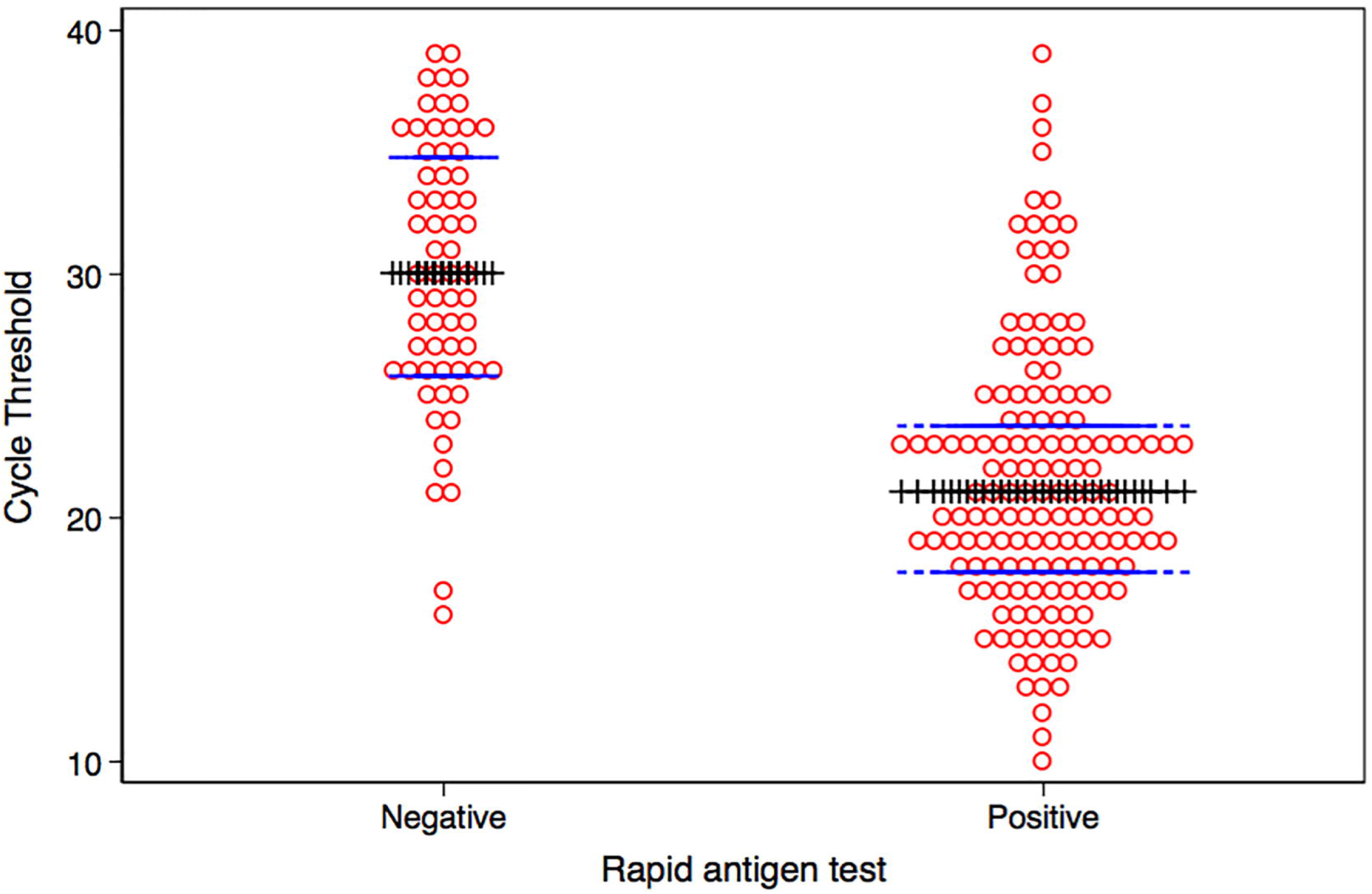
Mean sensitivity of the rapid test as a function of Cycle Thresholds (Ct) of the RT-PCR and 95% CI error bars. Sensitivity of the rapid test decreases with Ct values, indicating lower viral loads.

**FIGURE 2:**
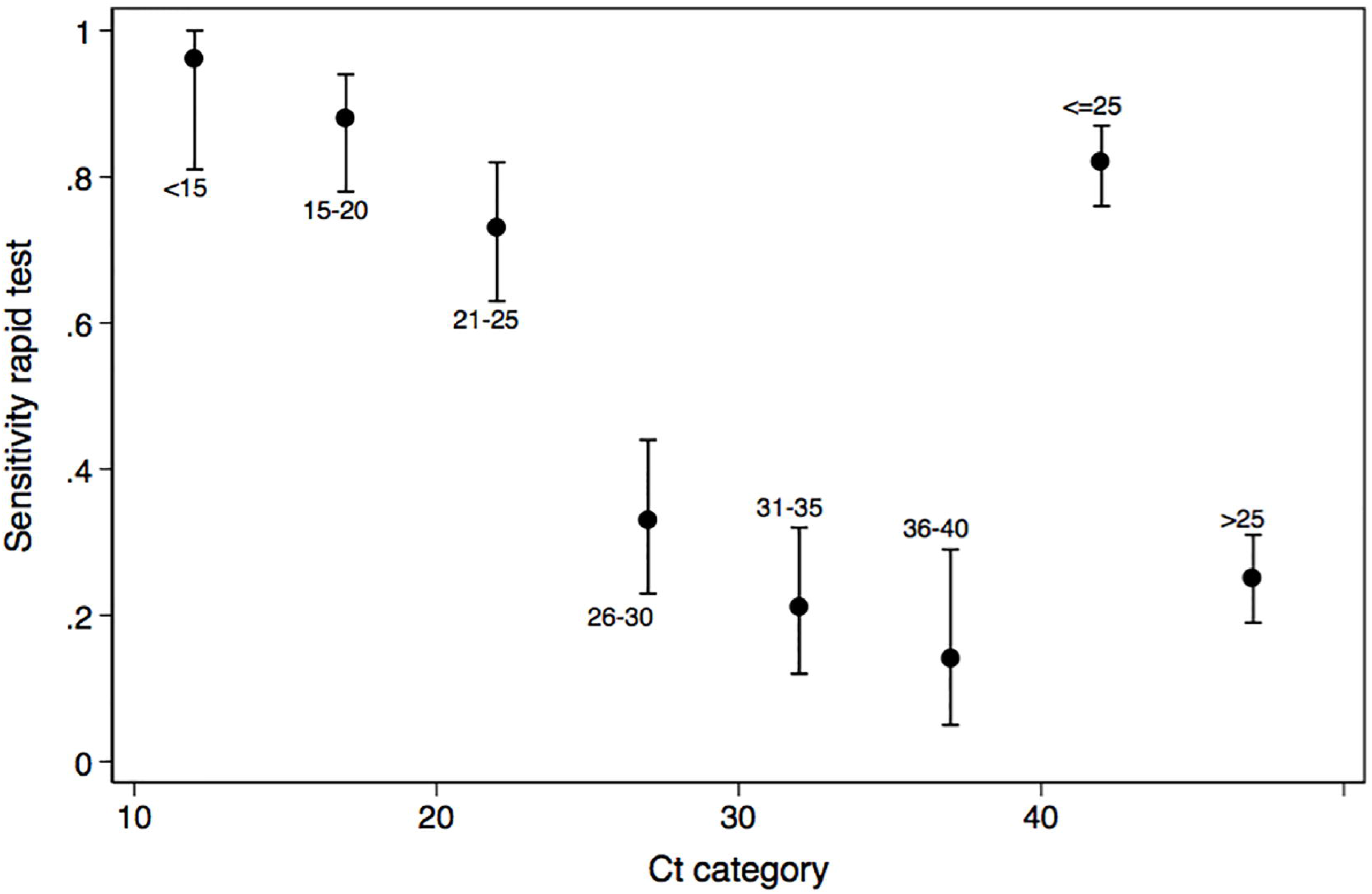
Mean sensitivity of the rapid test as a function of the duration of respiratory symptoms in participants positive for the RT-PCR, and 95%CI error bars. Sensitivity drops significantly with duration of symptoms.

**FIGURE 3:**
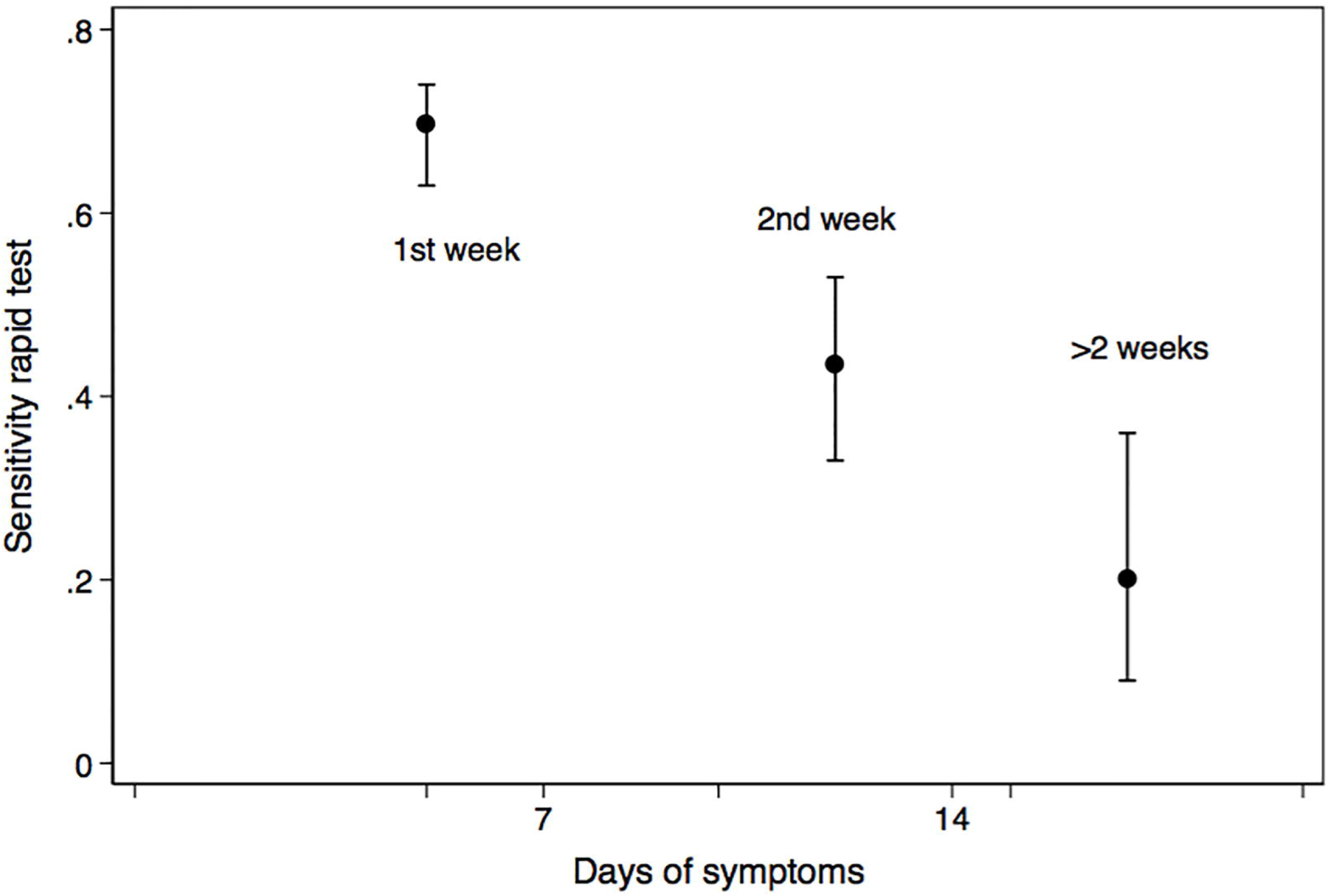
Dot graph of Ct (red circles) with median value, and 25th and 75% percentiles (blue lines), in individuals with normal (left side) or abnormal tests (right side). Those with positive rapid tests have lower Ct values.

**Table 4** depicts the diagnostic performance of the rapid test for the whole group and stratified by time of duration of symptoms. Overall sensitivity of the Panbio™ test was 54.4% (95%CI, 51-57) with a positive likelihood ratio of 35.7, a negative likelihood ratio of 0.46 and a Receiver-Operating Characteristics (ROC) curve area of 0.77. For participants presenting during the first week of symptoms, sensitivity was 69.6% (95%CI, 66-73), decreasing considerably with a longer duration of symptoms. In fact, the presence of symptoms was a predictor of positivity for the rapid test (sensitivity of 58.3% in participants with symptoms, compared with one of 26.3% in persons with no symptoms but with positive RT-PCR). On the other hand, specificity of the rapid test was above 97.8% in all groups.

**Table 4.**
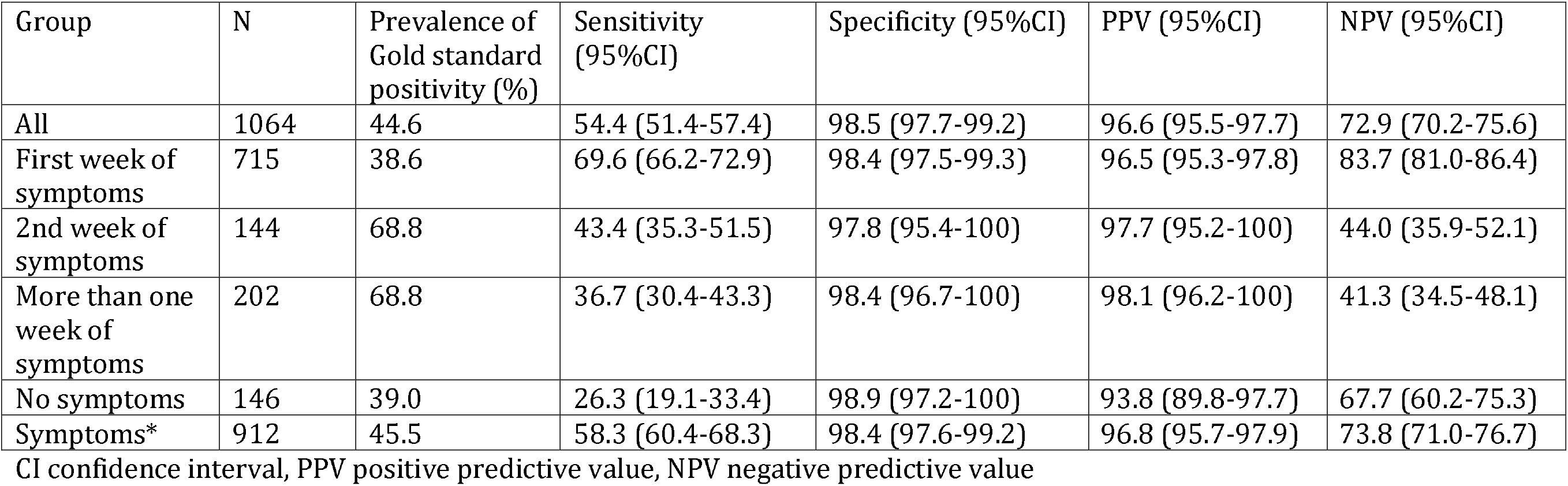
Diagnostic performance of the rapid antigen test.

Concordance between the rapid test and RT-PCR (Table S3) during the first week of symptoms was 0.71 (Standard Error, SE 0.04), but on considering all the participants it was 0.55 (SE 0.03). For participants presenting during the second week of symptoms, sensitivity was 0.31 (0.06) and 0.13 (SE 0.07) for participants with more than 2 weeks of symptoms.

A positive result on the RT-PCR was predicted by a positive result on the rapid test (OR 100; 95%CI; 48-208), by days of symptoms (OR 1.09; 95%CI, 1.05-1.11), male sex (OR, 1.5; 95%CI, 1.05-2.11), and age (OR, 1.01; 95%CI, 1.00-1.02) in a logistic regression model with a Pseudo R^2^ of 0.38, and these same variables were associated with the Ct.

In a multivariate logistic regression, on modeling positivity for the rapid test, the most important predictor was the RT-PCR test and especially the Ct (aOR, 0.79; 95%CI, 0.75-0.83), but also the days of symptoms (aOR, 0.90; 95%CI, 0.85-0.96) adjusting for age, sex, and comorbidities (Pseudo R^2^ 0.34).

The diagnostic characteristics of the rapid test varied across sites, with substantial heterogeneity in sensitivity (I^2^ 80.9; 95%CI, 68-93) and specificity (I^2^ 73.5; 95%CI, 54-92), with a relevant influence of centers contributing fewer participants (**Figure 4**).

**FIGURE 4:**
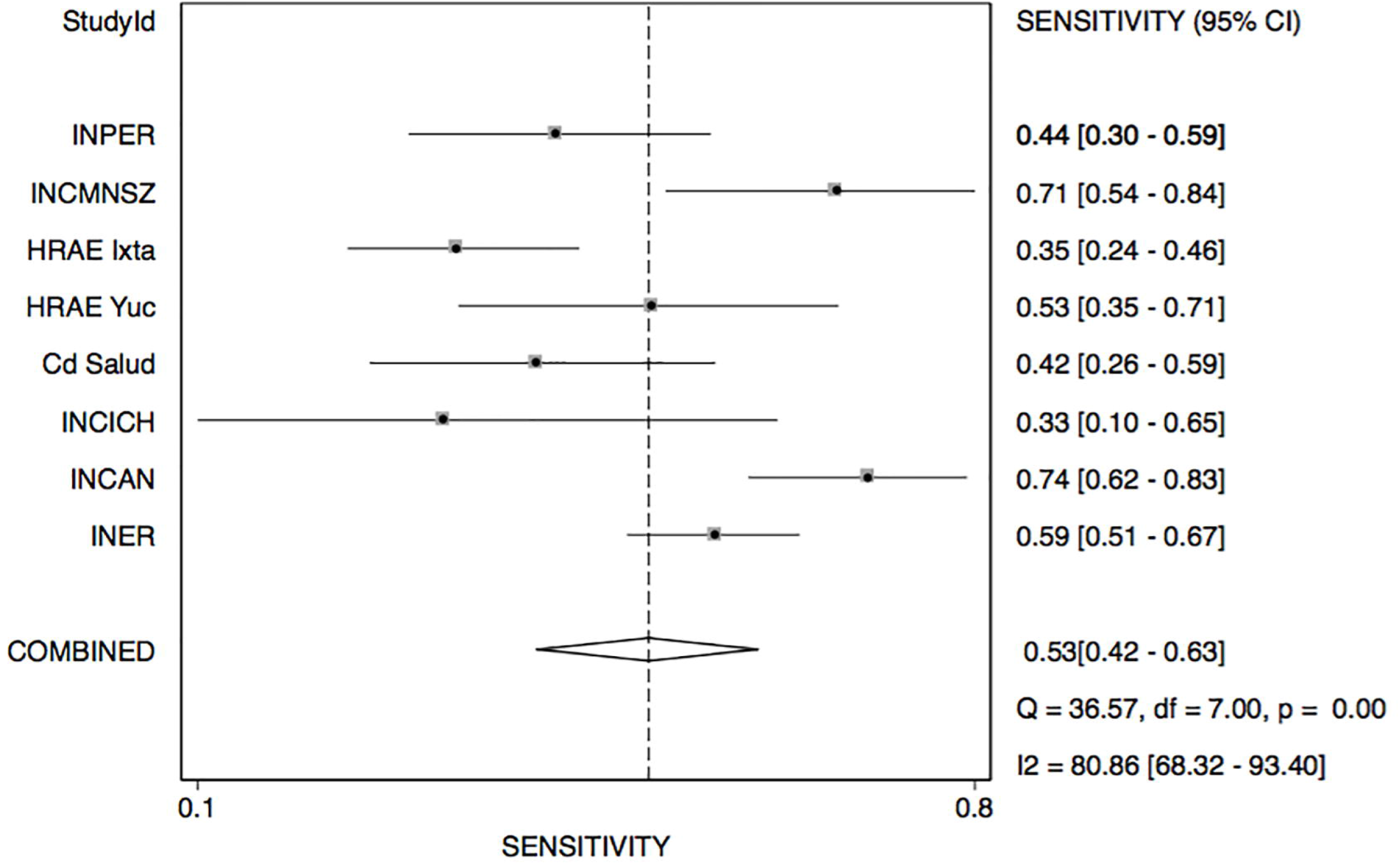
Forest plot depicting combined sensitivity and specificity (forest plot) as a function of categories of days of symptoms. Combined sensitivity was 0.53 (range 0.42-0.63) with I^2^ of 90.1% (range 68-83), whereas specificity was consistent and an overall value of 0.99 (0.93-1.0) with I^2^ of 73 (range 55-92)

## DISCUSSION

We present the results of a validation experiment of a commercial POC rapid antigen rapid test for SARS-CoV-2 infection, revealing high variability in sensitivity with respect to time from symptoms onset and to viral load. In our study, during the first week of symptoms, the sensitivity of the Panbio™ rapid antigen test was 69.6%, although the overall sensitivity was 54.4%, much lower than that reported by the manufacturer in the insert (11). The test was performed in patients with a longer duration of symptoms, or with no respiratory symptoms, predominantely contacts of symptomatic cases, often common in ER or at screening sites, especially during outbreak peaks when accesses to hospitals was difficult.

On the other hand, the specificity was consistently high in all subgroups.

In studies conducted elsewhere with the Abbott Panbio™ test, lower sensitivities have also been found, ranging from 45% in pediatric patients with less than 5 days of symptoms compatible with COVID-19 (15), 48% in household contacts of positive cases, 37% in non-domestic contacts (16), and up to 73% in primary-care patients (17). In the meta-analysis, days of evolution were strongly associated with the rapid test result, with a positivity of 86.5% if symptoms were present for fewer than 7 days and 54% if these were more than 7 days (17).

In a study conducted under normal working conditions, the sensitivity was 73% in the Netherlands and 81% in Aruba, and higher than 95% if the Ct was <32% (18). In all studies, the specificity has been 100% or close to 100%.

In another study in symptomatic patients in primary care and in routine work, sensitivity was 80% and specificity 100% (19).

The Canadian authorities issued recommendations that take into account sensitivities were than those incorporated into the annex to the test (20).

An update of a Cochrane review (21), including 78 studies (20 pre-prints) and 24,087 samples (about one third positive for SARS-CoV-2), confirmed a substantial variation in sensitivity according to the presence or absence of symptoms (72% vs. 58%), first v. second week of symptoms onset (78% vs. 51%), a Ct of ≤25 v. >25 (94.5 vs 40.7%) (21). Sensitivities reported for rapid antigen tests for SARS-CoV-2 from different manufacturers ranged from 34% to 88%, but specificities were in general above 99% (21). The pooled sensitivity of the Abbott Panbio™ test (11 studies) was 75.1% in symptomatic patients during the first week of symptoms, but dropped to 58% in asymptomatic individuals (21). Specificity was 99.5%. In the first week of symptoms, the sensitivity in our study was 69.6%, and specificity was 98.5%.

It Is relevant that only one study compiled by the recent Cochrane analysis (22) had a lower limit of confidence of test sensitivity of above 80%, the recommended cut-off point by the WHO for this type of tests.

In our analysis, we observed a strong association between the result of the rapid test and viral load, estimated with the Ct, with no positive results in samples with a Ct of >39. It is also relevant that the test was performed in a group of persons with >5 days from the onset of symptoms, a group in which the positivity decreased significantly due to an expected lower viral load. Nevertheless, even in this group in which sensitivity is considerably lower, a positive test would be highly informative.

It is noteworthy that, with symptoms of longer duration, the cost-effectiveness of applying POC rapid antigen tests would drop progressively. Thus, given that specificity is high, the greatest clinical advantage of utilizing a rapid antigen test would present when the result is positive, in which case, a confirmatory RT-PCR would not be needed.

In the presence of symptoms compatible with COVID-19, or in a period of a high incidence rate of infection in the community, a negative RT-PCR test result would be unreliable, even during the first week of symptoms. In these cases, the RT-PCR test is usually repeated a second, or even a third time (23). As the infection progresses, the viral and nucleic-acid load tends to decrease and the RT-PCR test tends to be negative, while antibody titers against SARS-CoV-2 begin to appear (24-27). Thus, confirming a SARS-CoV-2 infection ideally involves both the positivity of an RT-PCR test (not necessarily the first test), and the consideration of a combination of epidemiological variables, including the rate of community transmission, the presence of compatible symptoms and the presence of antibodies.

False/positive RT-PCR test results have been reported rarely, and are attributed to contamination with viral genetic material in any of the steps between sampling and processing.

On the other hand, in the case of a negative antigen test, confirmation with RT-PCR will be required, especially in the presence of compatible symptoms, a high rate of community infections, or in the case of persons with direct contact with confirmed COVID-19 patients. In any case, according to our observations, if a POC rapid antigen test were employed, 69.6% of the RT-PCR tests would be avoided during the first week of symptoms, which represents a considerable saving. The latter represents an enormous advantage in settings where decision-making is needed, and where the lack of infrastructure and high costs render it difficult to implement molecular testing.

Our study was observational, subject to all possible biases or cross-sectional studies, performed predominately during daytime, the shift with more available staff and in-training personnel. Testing was carried out with the Mexican health system and personnel under stress, a very different situation from that of rapid tests performed under controlled circumstances, in a laboratory, by the same expert personnel, and this can explain, at least in part. the reduced sensitivity. On the other hand, our results are expected to be closer to what can be observed under outbreaks that saturate or overwhelm the screening sites and ER, that is, more demanding circumstances than those found under strictly controlled laboratory testing. Furthermore, overall positivity in participating hospitals to the RT-PCR was quite high (44.6%, ranging from 27 to 93% in different hospitals) allowing for a proper evaluation of sensitivity and specificity. Real conditions of use may be even more demanding than those present in our study for example if testing includes primary care, community hospitals, during all shifts including weekends and nights, characterized by the presence of fewer personnel, and especially if the overall positivity of the tests or community transmission is low.

## CONCLUSIONS

We have shown that the sensitivity of a SARS-CoV-2 rapid antigen test is limited, with an overall estimated value of 70% in the present study. As expected, sensitivity was highly associated with time from the onset of symptoms and with viral load. The low sensitivity, and the high specificity observed in the test make it necessary to confirm all negative results, especially in the presence of symptoms compatible with COVID-19 and in settings with high community-infection rate. Nevertheless, the use of rapid antigen tests could be highly beneficial to screen for positive tests and to reduce the number of cases to be confirmed, permitting the making of quick clinical decisions.

## Supporting information

Positivity of the rapid test was strongly associated with Ct (a surrogate for viral load) (Figure 1), (Table S1)

Positivity also depended on the days since the onset of symptoms (Figure 3) (Table S2),

## Data Availability

Data and Code Availability
The data and codes related to the findings of this study will be available from the corresponding author after publication, upon reasonable request, especially from investigators compiling studies to of COVID-19. The group formed by the principal investigators will analyze the quality of the proposal and the security offered for the data. The patient-level data, without individual identifiers, will be shared after approval of the submitted proposal.

## ACKNOWLEDGMENTS

The antigen test kits were donated by the WHO / PAHO, and the Mexican Health Secretariat purchased additional tests of the same type for the hospital care of the patients.

No other additional support was available, and the expenses incurred were covered by the participating hospitals. Care of patients with COVID-19 was complimentary in all participating hospitals.

---

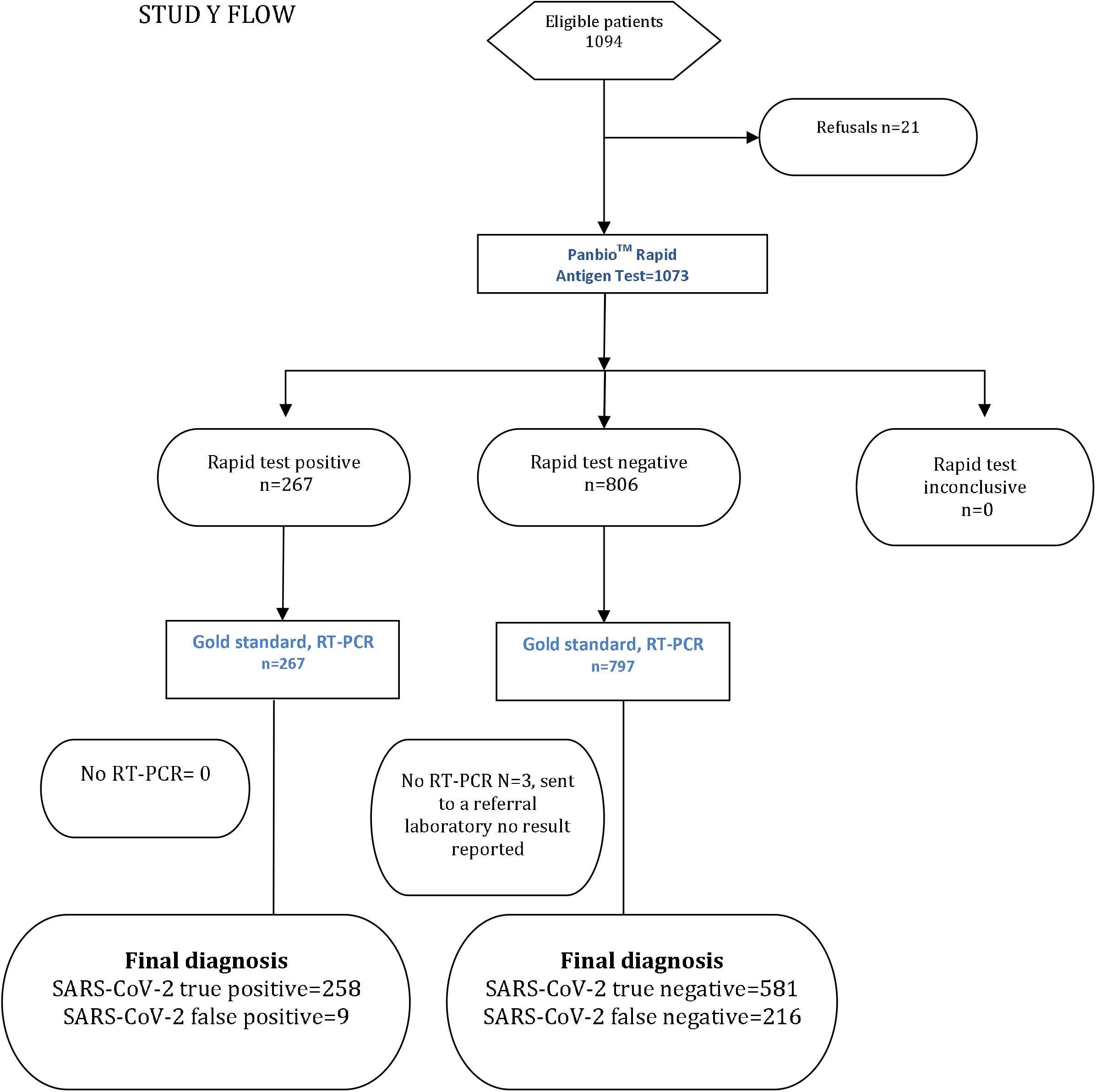

## REFERENCES

1. Benirschke RC, McElvania E, Thomson RB, Kaul KL, Das S. Clinical impact of rapid point-of-care pcr influenza testing in an urgent care setting: A single-center study. J Clin Microbiol 2019;57.

2. Rahamat-Langendoen J, Groenewoud H, Kuijpers J, Melchers WJG, van der Wilt GJ. Impact of molecular point-of-care testing on clinical management and in-hospital costs of patients suspected of influenza or rsv infection: A modeling study. J Med Virol 2019;91:1408–1414.

3. Shengchen D, Gu X, Fan G, Sun R, Wang Y, Yu D, Li H, Zhou F, Xiong Z, Lu B, Zhu G, Cao B. Evaluation of a molecular point-of-care testing for viral and atypical pathogens on intravenous antibiotic duration in hospitalized adults with lower respiratory tract infection: A randomized clinical trial. Clin Microbiol Infect 2019;25:1415–1421.

4. Wabe N, Li L, Lindeman R, Yimsung R, Dahm MR, McLennan S, Clezy K, Westbrook JI, Georgiou A. Impact of rapid molecular diagnostic testing of respiratory viruses on outcomes of adults hospitalized with respiratory illness: A multicenter quasi-experimental study. J Clin Microbiol 2019;57.

5. Schechter-Perkins EM, Mitchell PM, Nelson KP, Liu JH, Shannon A, Ahern J, Orr B, Miller NS. Point-of-care influenza testing does not significantly shorten time to disposition among patients with an influenza-like illness. Am J Emerg Med 2019;37:873–878.

6. Watson J, Whiting PF, Brush JE. Interpreting a covid-19 test result. Bmj 2020;369:m1808.

7. Dinnes J, Deeks JJ, Adriano A, Berhane S, Davenport C, Dittrich S, Emperador D, Takwoingi Y, Cunningham J, Beese S, Dretzke J, Ferrante di Ruffano L, Harris IM, Price MJ, Taylor-Phillips S, Hooft L, Leeflang MM, Spijker R, Van den Bruel A. Rapid, point-of-care antigen and molecular-based tests for diagnosis of sars-cov-2 infection. Cochrane Database Syst Rev 2020;8:Cd013705.

8. Kubina R, Dziedzic A. Molecular and serological tests for COVID-19 a comparative review of sars-cov-2 coronavirus laboratory and point-of-care diagnostics. Diagnostics (Basel) 2020;10.

9. Mina MJ, Parker R, Larremore DB. Rethinking covid-19 test sensitivity - a strategy for containment. The New England journal of medicine 2020.

10. Corman VM, Landt O, Kaiser M, Molenkamp R, Meijer A, Chu DK, Bleicker T, Brünink S, Schneider J, Schmidt ML, Mulders DG, Haagmans BL, van der Veer B, van den Brink S, Wijsman L, Goderski G, Romette J-L, Ellis J, Zambon M, Peiris M, Goossens H, Reusken C, Koopmans MP, Drosten C. Detection of 2019 novel coronavirus (2019-ncov) by real-time rt-pcr. Eurosurveillance 2020;25:2000045.

11. Abbot. Inser to Abbot Panbio COVID-19 ag rapid test device. 2020.

12. World Health Organization. A minimal common outcome measure set for COVID-19 clinical research. Lancet Infect Dis 2020;20:e192–e197.

13. Bujang MA. Requirements for minimum sample size for sensitivity and specificity analysis. Journal of Clinical and Diagnostic Research 2016.

14. Hajian-Tilaki K. Sample size estimation in diagnostic test studies of biomedical informatics. Journal of Biomedical Informatics 2014;48:193–204.

15. Villaverde S, Domínguez-Rodríguez S, Sabrido G, Pérez-Jorge C, Plata M, Romero MP, Grasa CD, Jiménez AB, Heras E, Broncano A, Núñez MDM, Illán M, Merino P, Soto B, Molina-Arana D, Bermejo A, Mendoza P, Gijón M, Pérez-Moneo B, Moraleda C, Tagarro A. Diagnostic accuracy of the panbio severe acute respiratory syndrome coronavirus 2 antigen rapid test compared with reverse-transcriptase polymerase chain reaction testing of nasopharyngeal samples in the pediatric population. J Pediatr 2021.

16. Torres I, Poujois S, Albert E, Colomina J, Navarro D. Evaluation of a rapid antigen test (panbio™ COVID-19 ag rapid test device) for sars-cov-2 detection in asymptomatic close contacts of COVID-19 patients. Clin Microbiol Infect 2021;27:636.e631-634.

17. Linares M, Pérez-Tanoira R, Carrero A, Romanyk J, Pérez-García F, Gómez-Herruz P, Arroyo T, Cuadros J. Panbio antigen rapid test is reliable to diagnose sars-cov-2 infection in the first 7 days after the onset of symptoms. J Clin Virol 2020;133:104659.

18. Gremmels H, Winkel BMF, Schuurman R, Rosingh A, Rigter NAM, Rodriguez O, Ubijaan J, Wensing AMJ, Bonten MJM, Hofstra LM. Real-life validation of the panbio™ COVID-19 antigen rapid test (abbott) in community-dwelling subjects with symptoms of potential sars-cov-2 infection. EClinicalMedicine 2021;31:100677.

19. Albert E, Torres I, Bueno F, Huntley D, Molla E, Fernández-Fuentes M, Martínez M, Poujois S, Forqué L, Valdivia A, Solano de la Asunción C, Ferrer J, Colomina J, Navarro D. Field evaluation of a rapid antigen test (panbio™ COVID-19 ag rapid test device) for COVID-19 diagnosis in primary healthcare centres. Clin Microbiol Infect 2021;27:472.e477-472.e410.

20. Interim guidance on the use of the abbott panbio™ COVID-19 antigen rapid test. Can Commun Dis Rep 2021;47:17–22.

21. Dinnes J, Deeks JJ, Berhane S, Taylor M, Adriano A, Davenport C, Dittrich S, Emperador D, Takwoingi Y, Cunningham J, Beese S, Domen J, Dretzke J, Ferrante di Ruffano L, Harris IM, Price MJ, Taylor-Phillips S, Hooft L, Leeflang MM, McInnes MD, Spijker R, Van den Bruel A, Cochrane C-DTAG. Rapid, point-of-care antigen and molecular-based tests for diagnosis of sars-cov-2 infection. Cochrane Database Syst Rev 2021;3.

22. Alemany A, Baró B, Ouchi D, Rodó P, Ubals M, Corbacho-Monné M, Vergara-Alert J, Rodon J, Segalés J, Esteban C, Fernández G, Ruiz L, Bassat Q, Clotet B, Ara J, Vall-Mayans M, G-Beiras C, Blanco I, Mitjà O. Analytical and clinical performance of the panbio COVID-19 antigen-detecting rapid diagnostic test. J Infect 2021.

23. Ramdas K, Darzi A, Jain S. ‘Test, re-test, re-test’: Using inaccurate tests to greatly increase the accuracy of COVID-19 testing. Nat Med 2020;26:810–811.

24. Ghaffari A, Meurant R, Ardakani A. COVID-19 serological tests: How well do they actually perform? Diagnostics (Basel) 2020;10.

25. Kilic T, Weissleder R, Lee H. Molecular and immunological diagnostic tests of COVID-19: Current status and challenges. iScience 2020;23:101406.

26. Laureano AFS, Riboldi M. The different tests for the diagnosis of COVID-19 - a review in brazil so far. JBRA Assist Reprod 2020;24:340–346.

27. Shyu D, Dorroh J, Holtmeyer C, Ritter D, Upendran A, Kannan R, Dandachi D, Rojas-Moreno C, Whitt SP, Regunath H. Laboratory tests for COVID-19: A review of peer-reviewed publications and implications for clinical use. Mo Med 2020;117:184-195.

