## Supplementary figures and images for "EVALUATION OF A RAPID ANTIGEN TEST FOR SARS-COV-2 IN SYMPTOMATIC PATIENTS AND THEIR CONTACTS: A MULTICENTER STUDY"

### Positivity also depended on the days since the onset of symptoms (Figure 3) (Table S2),

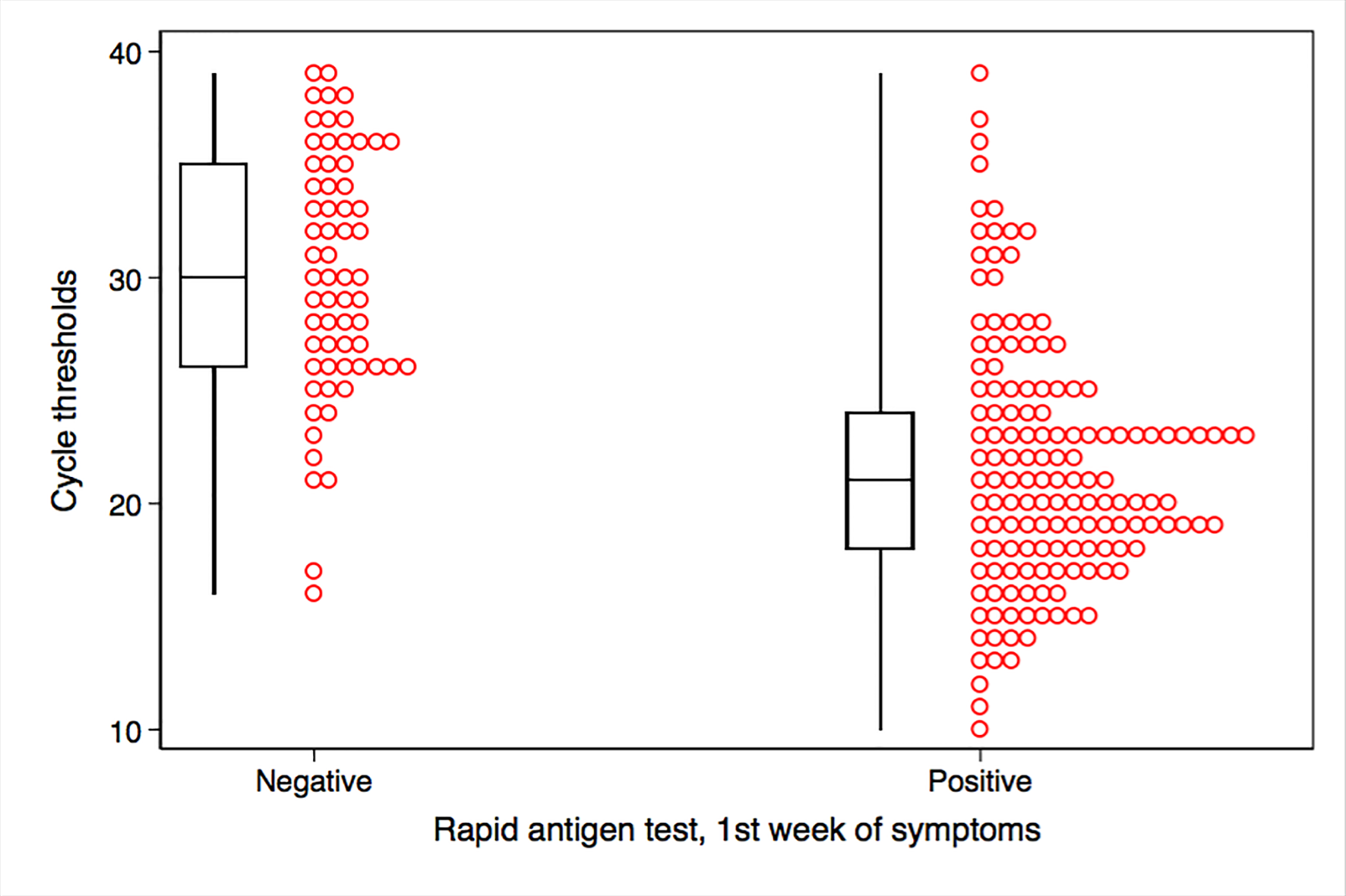

### Positivity of the rapid test was strongly associated with Ct (a surrogate for viral load) (Figure 1), (Table S1)

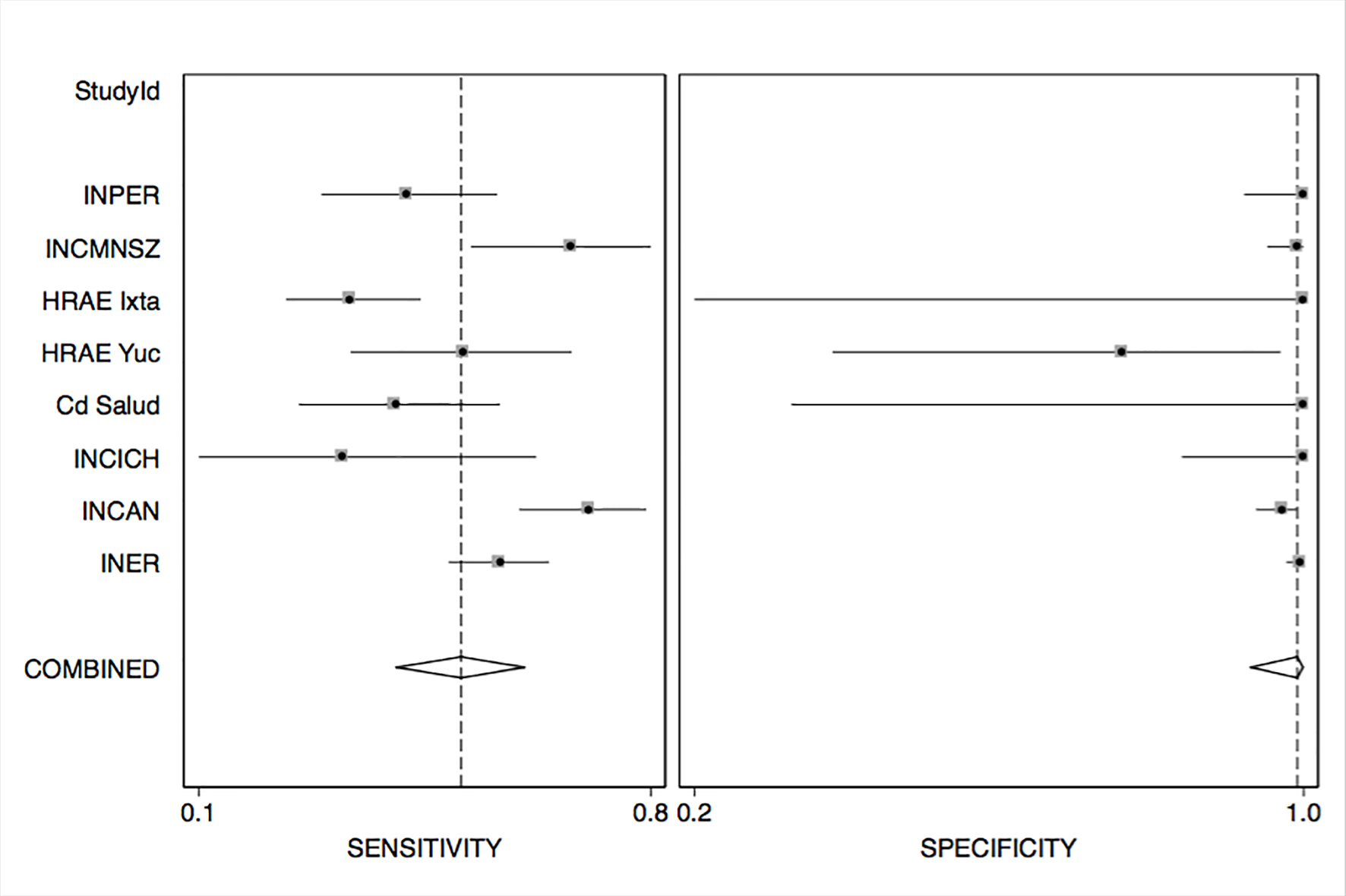
