## Supplementary material for "EVALUATION OF A RAPID ANTIGEN TEST FOR SARS-COV-2 IN SYMPTOMATIC PATIENTS AND THEIR CONTACTS: A MULTICENTER STUDY": ONLINE SUPPLEMENT

**TABLE S1: SENSITIVITY OF RAPID TEST ACCORDING TO CYCLE TRESHOLDS**

| Cycles to threshold | N | Antigen test positive | Sensitivity | 95%CI lower bound | 95% CI higher bound |
| --- | --- | --- | --- | --- | --- |
| <15 | 27 | 26 | 0.96 | 0.81 | 1.0 |
| 15-20 | 75 | 66 | 0.88 | 0.78 | 0.94 |
| 21-25 | 92 | 67 | 0.73 | 0.63 | 0.82 |
| 26-30 | 88 | 29 | 0.33 | 0.23 | 0.44 |
| 31-35 | 69 | 14 | 0.21 | 0.12 | 0.32 |
| 38-39 | 37 | 5 | 0.14 | 0.05 | 0.29 |
| ct<=25 | 194 | 159 | 0.82 | 0.76 | 0.87 |
| ct>25 | 195 | 48 | 0.25 | 0.19 | 0.31 |
| All subjects with Ct reported | 389 | 207 | 0.54 | 0.48 | 0.58 |

**TABLE S2: SENSITIVITY OF THE RAPID ANTIGEN TEST ACCORDING TO DAYS OF SYMPTOMS (in subjects positive to** RT-PCR)

| Days of symptoms | N | Positive rapid test | Rapid test Sensitivity | 95% CI lower bound | 95%CI upper bound |
| --- | --- | --- | --- | --- | --- |
| First week | 278 | 192 | 0.70 | 0.63 | 0.74 |
| 2nd week | 100 | 43 | 0.43 | 0.33 | 0.53 |
| >2 weeks | 40 | 8 | 0.20 | 0.09 | 0.36 |

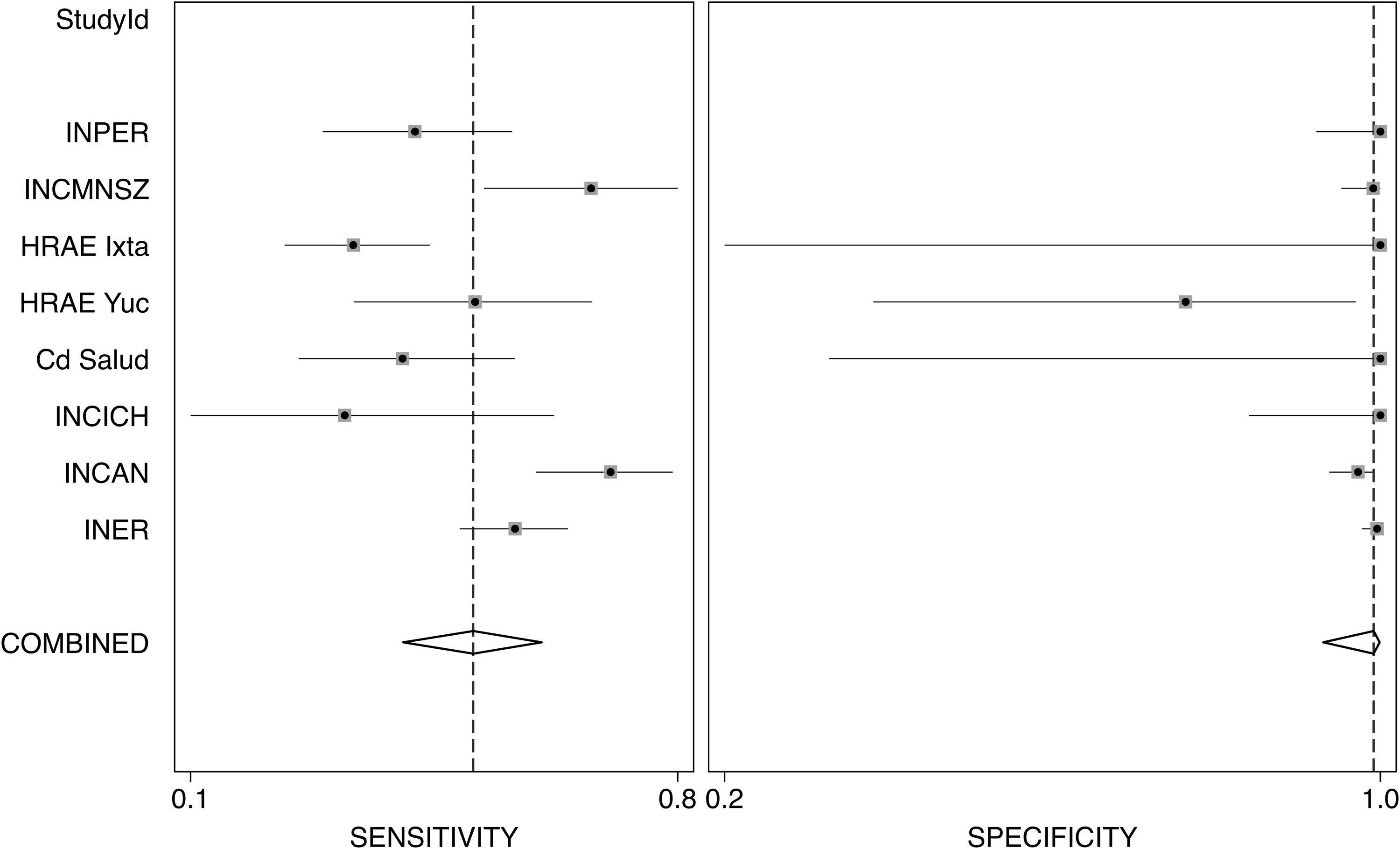

FIGURE S1- FOREST PLOT PER HOSPITAL PARTICIPATING INCLUDING SENSITIVITY AND SPECIFICITY

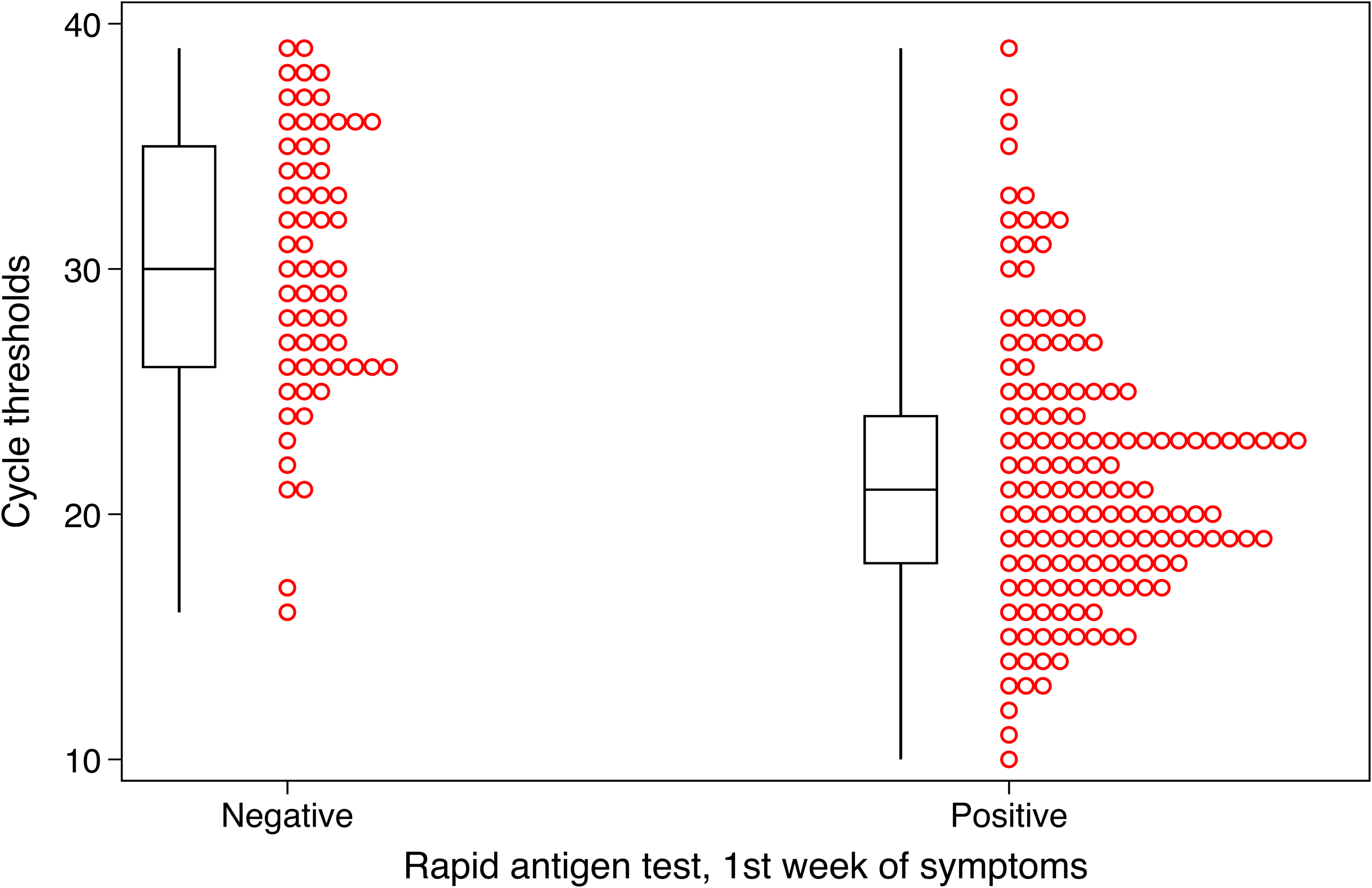

FIGURE S2 BOX AND DOT PLOT OF Ct´S ACCORDING TO POSITIVITY OF THE RAPID TEST

Table S3: Agreement between rapid antigen test (Panbio) and RT-PCR.

| Group of patients | Agreement | Expected agreement | Kappa | Kappa SE | P |
| --- | --- | --- | --- | --- | --- |
| All participants | 0.78 | 0.52 | 0.55 | 0.03 | <0.0001 |
| First week of symptoms | 0.87 | 0.54 | **0.71** | 0.04 | <0.0001 |
| 2nd week of symptoms | 0.60 | 0.42 | 0.31 | 0.06 | <0.0001 |
| >2 weeks of symptoms | 0.45 | 0.36 | 0.13 | 0.07 | 0.02 |

SE= standard
